# Development of the reproduction number from coronavirus SARS-CoV-2 case data in Germany and implications for political measures

**DOI:** 10.1101/2020.04.04.20053637

**Authors:** Sahamoddin Khailaie, Tanmay Mitra, Arnab Bandyophadhyay, Marta Schips, Pietro Mascheroni, Patrizio Vanella, Berit Lange, Sebastian C. Binder, Michael Meyer-Hermann

## Abstract

The novel Coronavirus SARS-CoV-2 (COVID-19) has induced a world-wide pandemic and subsequent non-pharmaceutical interventions (NPI) in order to control the spreading of the virus. NPIs are considered to be critical in order to at least delay the peak number of infected individuals and to prevent the health care system becoming overwhelmed by the number of patients to treat in hospitals or in intensive care units (ICUs). However, there is also increasing concern that the NPIs in place would increase mortality because of other diseases, increase the frequency of suicide and increase the risk of an economic recession with unforeseeable implications. It is therefore instrumental to evaluate the necessity of NPIs and to monitor the progress of containment of virus spreading.

We used a data-driven estimation of the evolution of the reproduction number for viral spreading in Germany as well as in all its federal states. Based on an extended infection-epidemic model, parameterized with data from the Robert Koch Institute and, alternatively, with parameters stemming from a fit to the initial phase of COVID-19 spreading in different regions of Italy, we found that the reproduction number was decreased to a range below 1 in all federal states. The development in Germany suggests that NPIs can be partially released based on an established new culture of social distancing, face masks and mutual care within the population. However, any release of measures delays reaching low incidence numbers. The strategy to reduce daily new cases to a sufficiently low level to be controlled by contact tracing and testing turned out to work in Germany. This requires a responsible behaviour of the population, optimised contact tracing techniques and extended testing capacities in contact clusters.

**Author summary:** As of mid-June Germany was able to control the pandemic to an extent that the health care system was not overwhelmed and the daily new reported infections appear under control. We analysed the evolution of the reproduction number during the epidemic in Germany and the efficiency of non-pharmaceutical interventions (NPIs) in containing viral spread. The results suggest that the cultural change induced by NPIs in interpersonal interactions and distancing allows for a partial release of political measures. However, this requires a functional case isolation and contact tracing of new cases by local health departments, early identification of contact clusters and consequent isolation of those clusters.

## Introduction

The outbreak of the novel Coronavirus SARS-CoV-2 (CoV) in China has induced a world-wide pandemic. The comparably high lethality in the elderly population and in patients with comorbidities [1], together with a widely absent immunisation of the population against the novel virus as well as the limited health system capacity estimated to become overwhelmed by an unlimited virus spreading [2], led to non-pharmaceutical interventions (NPIs) to reduce virus transmission mostly by reducing inter-individual contacts. The aim of these measures was to achieve at least a delay of viral spreading, allowing the health care system to extend its capacities and to treat less patients per time or, ideally, achieve a complete stop of viral spreading. The NPIs installed in Germany have been effective in containing viral dissemination [3]. Hence, in the light of economic damage incurred by restrictions [4], a gradual release of NPIs was decided with moderate effects on virus transmission. However, the delay in the range of the serial interval, which is in the range of 4.5 days for CoV [5, 6], and an inevitable delay in testing and reporting implies that any sudden outbreaks may be recognized too late and careful continuous monitoring of the infection dynamics on a regional basis is required. Thus, current political decisions need foundational information about current infection dynamics and their response to changes in NPIs such as partial release of contact restrictions or school openings, ideally on a regional basis. In fact, a declining or stable number of daily reported cases despite releasing measures can be misleading if the present trend of the achieved reproduction number, the delay between changes in the infection dynamics and their manifestations in reported case numbers are not taken into account. Furthermore, the high variance of the locally reported new cases adds to this uncertainty. Thus, it is extremely important to construct a model that captures not only the disease dynamics but also has the potential to provide information on the trend of the outbreak by considering the time-dependence of the reproduction number for COVID-19. The situation in Europe was recently analysed [6]. Here, a systematic analysis of the development of the reproduction number over the time period of the COVID-19 outbreak in Germany and in all federal states of Germany is provided.

A second level of information necessary for political decisions on NPIs is the prospective development of the outbreak under different scenarios. A too early release of NPIs risks to abandon the current level of containment and to initiate a new wave of viral spreading [7]. A too long application of NPIs carries the risk of collateral damage and imposes a strong economic burden [4]. In view of the currently achieved reproduction number in Germany and its federal states, a partial release of NPIs was decided, including partial school re-openings and resumption of catering and hotel business under certain restrictions, including mask obligations in public. The effects of such re-openings are hard to predict and require careful monitoring of local infection parameters and their implications for forecasting the immediate future development of the infection. Our contribution offers predictions based on current developments in terms of infections, number of hospital beds and intensive care units (ICUs) needed to treat patients with severe disease progression, and fatalities. This analysis provides additional information on when and how strongly to react to potential infection waves in order to avoid unacceptably high mortality and morbidity as well as excessive demands on the health care system.

The aim of this work is a quantitative evaluation of the reproduction number under the influence of NPIs in Germany and its federal states, together with a prospective estimation of the outbreak given a release of NPIs with uncertain consequences that may lead to a moderate or higher increase in new infections. As this information needs to be up-to-date for the purpose of closely monitoring effects of policy changes, we provide daily updates of our analysis results online [8].

## Materials and methods

The implemented SECIR (Susceptible-Exposed-Carrier-Infected-Recovered)-model is a deterministic ordinary-differential-equation model adapted to the specific properties of SARS-CoV-2 viral infections. It distinguishes healthy individuals without immune memory of COVID-19 (*S*), infected individuals without symptoms but not yet infectious (*E*), infected individuals without symptoms who are infectious (*C*_*I,R*_), and detected (*I*_*H,R*_) and undetected (*I*_*X*_) symptomatic patients. Further, compartments for hospitalization (*H*_*U,R*_) and intensive care units (*U*_*D,R*_) were introduced to monitor the load on the healthcare system. Detected patients recover from different states of the disease (*R*_*Z*_) or die (*D*). Undetected individuals who went through the infection are monitored (*R*_*X*_). The quantities are defined and the model is summarized in Fig. 1 with parameters in Table 1. The model equations read

**Table 1.** Parameter sets of the SECIR model: determination of the boundaries for literature-based parameter set was based on the interpretation of the values given in the references and are discussed in the supporting information. The Italy-based parameter set was determined by fitting the data for different regions of Italy and providing minimum and maximum over all regions.

| Parameter | References | Parameter set from literature |  | Parameter set from fitting Italy data |  |
| --- | --- | --- | --- | --- | --- |
|  |  | Min | Max | Min | Max |
| $R_1$ | Variable; fitted | | | | |
| $R_2$ | $1/R_2 = 5.2 - 1/R_3$ ; median incubation period is 5.2 days [13] | | | | |
| $R_3$ | [5, 14] | $\frac{1}{4.2}$ | $\frac{2}{5.2}$ | 0.2381 | 0.2382 |
| $R_4$ | [15] | $\frac{1}{14}$ | $\frac{1}{4}$ | 0.0714 | 0.25 |
| $R_5$ | [16, 17] | $\frac{1}{16}$ | $\frac{1}{7}$ | 0.0625 | 0.105 |
| $R_6$ | [18, 19] | $\frac{1}{7}$ | $\frac{1}{2.5}$ | 0.4 | 0.4 |
| $R_7$ | [12, 16] | $\frac{1}{14}$ | $\frac{1}{4}$ | 0.2857 | 1 |
| $R_8$ | [17] | $\frac{1}{16}$ | $\frac{1}{5}$ | 0.0625 | 0.0625 |
| $R_9$ | $\frac{1}{R_9} = \frac{1}{R_3} + \left(0.5 \times \frac{1}{R_4}\right)$ | | | | |
| $R_{10}$ | [20] | $\frac{1}{7.5}$ | $\frac{1}{3.5}$ | 0.1333 | 0.2857 |
| $\delta_0$ | [17, 21] | 0.15 | 0.77 | 0.15 | 0.77 |
| $\alpha$ | [1] | 0.01 | 0.5 | 0.01 | 0.0870 |
| $\beta$ | Assumed | 0.05 | 1 | 0.05 | 0.1 |
| $\rho_0$ | [21, 22] | 0.10 | 0.35 | 0.35 | 0.35 |
| $\vartheta_0$ | [16, 21] | 0.15 | 0.4 | 0.15 | 0.4 |
| $\mu$ | Assumed; varied | | | | |

**Fig 1.**
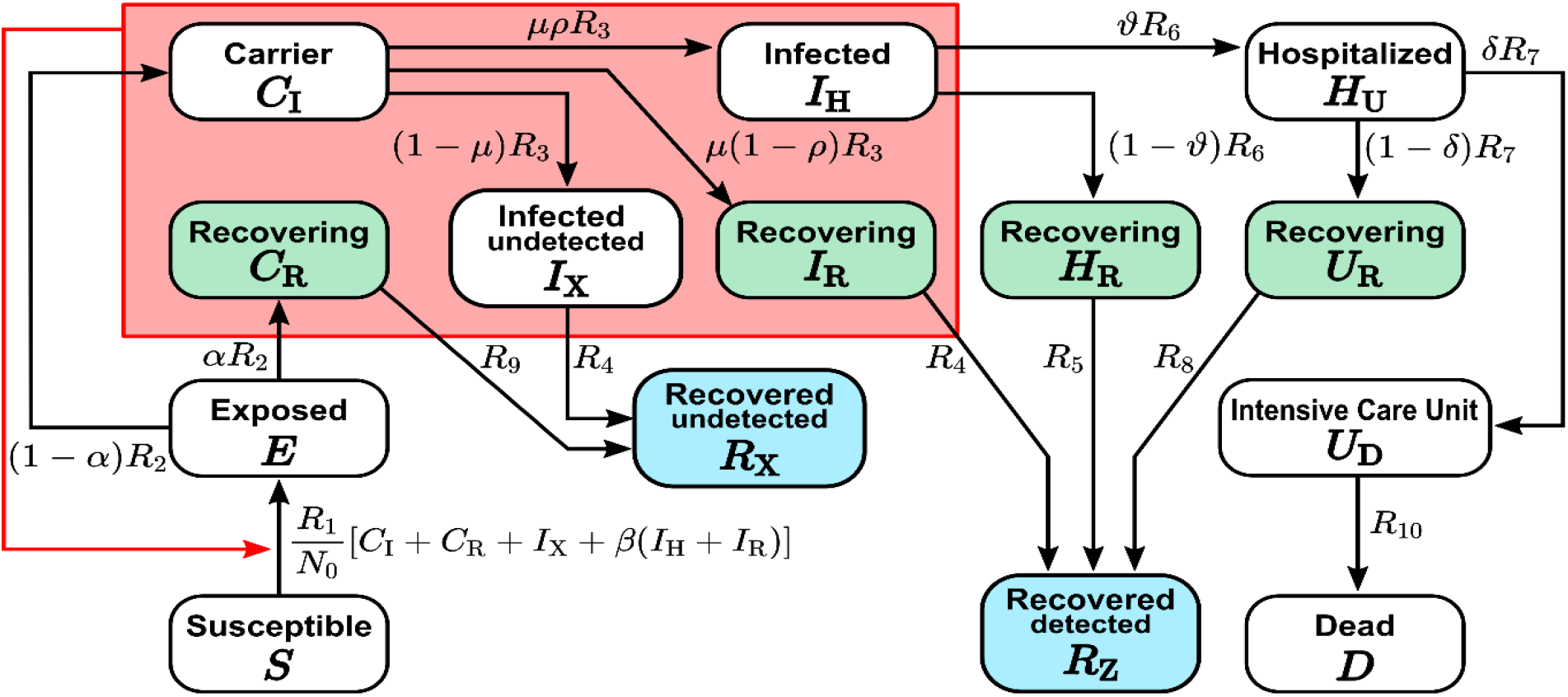
The scheme of the SECIR model, which distinguishes susceptible (*S*), healthy individuals without immune memory of CoV, exposed (*E*), who already carry the virus but are not yet infectious to others, carriers (*C*_*I,R*_), who carry the virus and are infectious to others but do not yet show symptoms, infected (*I*_*H,R,X*_), who carry the virus with symptoms and are infectious to others, hospitalized (*H*_*U,R*_), who experience a severe development of the disease, transferred to intensive care unit (*U*_*R,D*_), dead (*D*), and recovered (*R*_*D,X*_), who acquired immune memory and cannot be infected again. Recovery happens from each of the states *C*_*R*_, *I*_*X*_, *I*_*R*_, *H*_*R*_, *U*_*R*_. See Table 1 for parameter values.

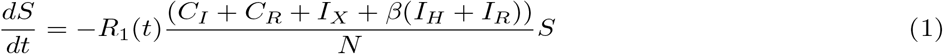

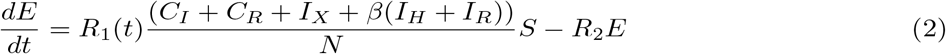

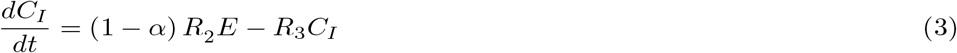

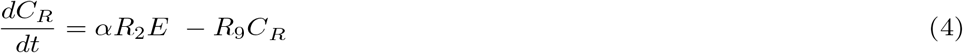

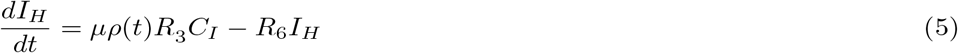

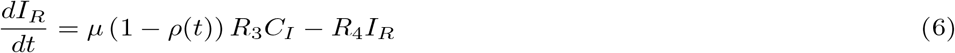

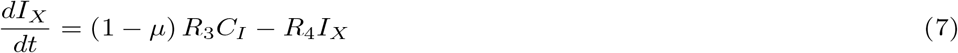

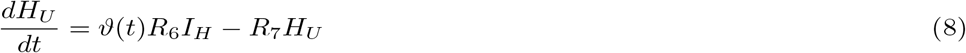

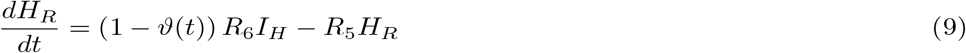

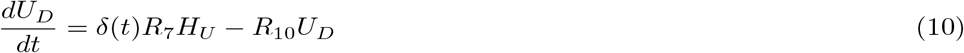

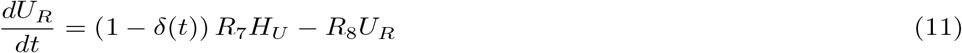

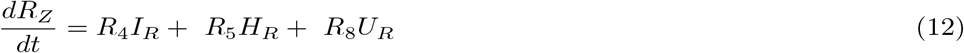

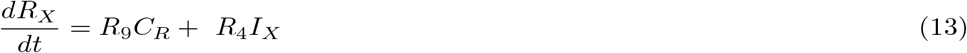

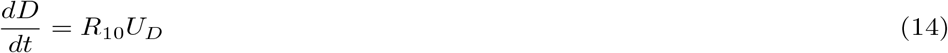

The rates *R*_2,…,10_ denote the inverse time of transition between the respective states and can be estimated from literature. Parameter *R*_1_ is fitted to the course of reported case numbers in a sliding time window, and therefore, is a time-varying parameter. Greek letters denote fractions of individuals with a particular fate. Parameters *ρ, ϑ* and *δ* have a time-varying component modeled with a logistic function

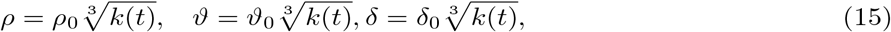

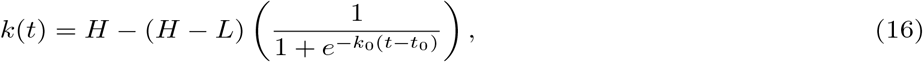

where *t* corresponds to the time after the starting of the epidemic and *H* = 0.156, *L* = 0.011, *k*_0_ = 0.25, and *t*_0_ = 61 are obtained from fitting the case fatality rate (CFR = *ρϑδ*), which changed over the course of the epidemic in Germany. This is due to changing testing frequencies [9] and the shifting age structure of the infected over time [10]; therefore, the CFR estimate is not reflecting a real change in the fatality rate of the virus.

The model further distinguishes recovered cases according to whether they were detected or not. While the age distribution can be included in such models [11], our conclusions turned out insensitive to differences in demographics, such that we decided to neglect age-specific parameters. These differences may be more important for the analysis of smaller districts; however, case numbers in smaller districts might not be sufficient for a proper discrimination of age groups.

### Parameterisation

The model parameters are critical for the overall behaviour of the model and for the quality of the predictions derived from it. For the sake of robustness of the results, we followed two different strategies on how to determine the model parameters. The analysis presented below was repeated with the parameter sets derived from both strategies.

The first strategy was based on literature research (e.g. [12]), where estimates of most of the model parameters were found (see Table 1). The uncertainty of the parameters were used to determine ranges of possible values for each parameter (Table 1). These ranges were subsequently used to determine the distribution of *R*_*t*_ values.

In the second approach, we kept model parameters open and fitted them to the cases reported in different regions of Italy until March 24^th^, 2020. Thus, we include the initial phase of the outbreak with comparably low effects from the overwhelmed health care system. As data for cumulative infected, hospitalized, ICU, deaths, and recovered are available (Italy Data on Coronavirus 2020), the fitting was able to substantially narrow down the parameter space. Fitting was repeated for each region in Italy separately. The diversity of parameter values determined the ranges of parameter variation (Fig. 2(A)) used to determine the distribution of *R*_*t*_ values (Fig. 3).

**Fig 2.**
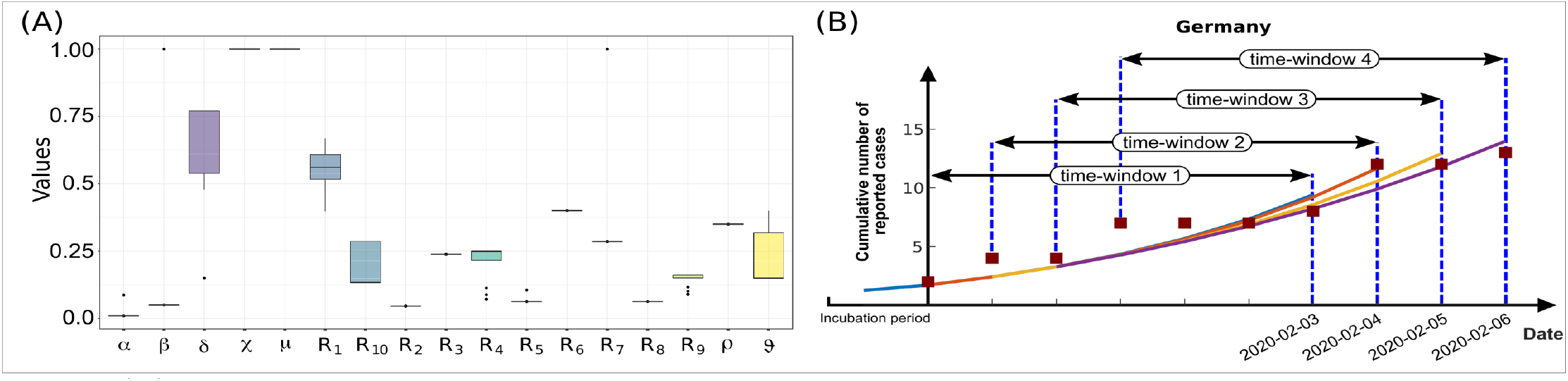
(A) Variability of parameters fitted to the number of reported, hospitalized, ICU, dead and recovered cases in different regions of Italy. Table 1 recollects the parameter ranges. (B) Scheme of the shifting time window and repeated fitting to the time course of the reported cases data.

**Fig 3.**
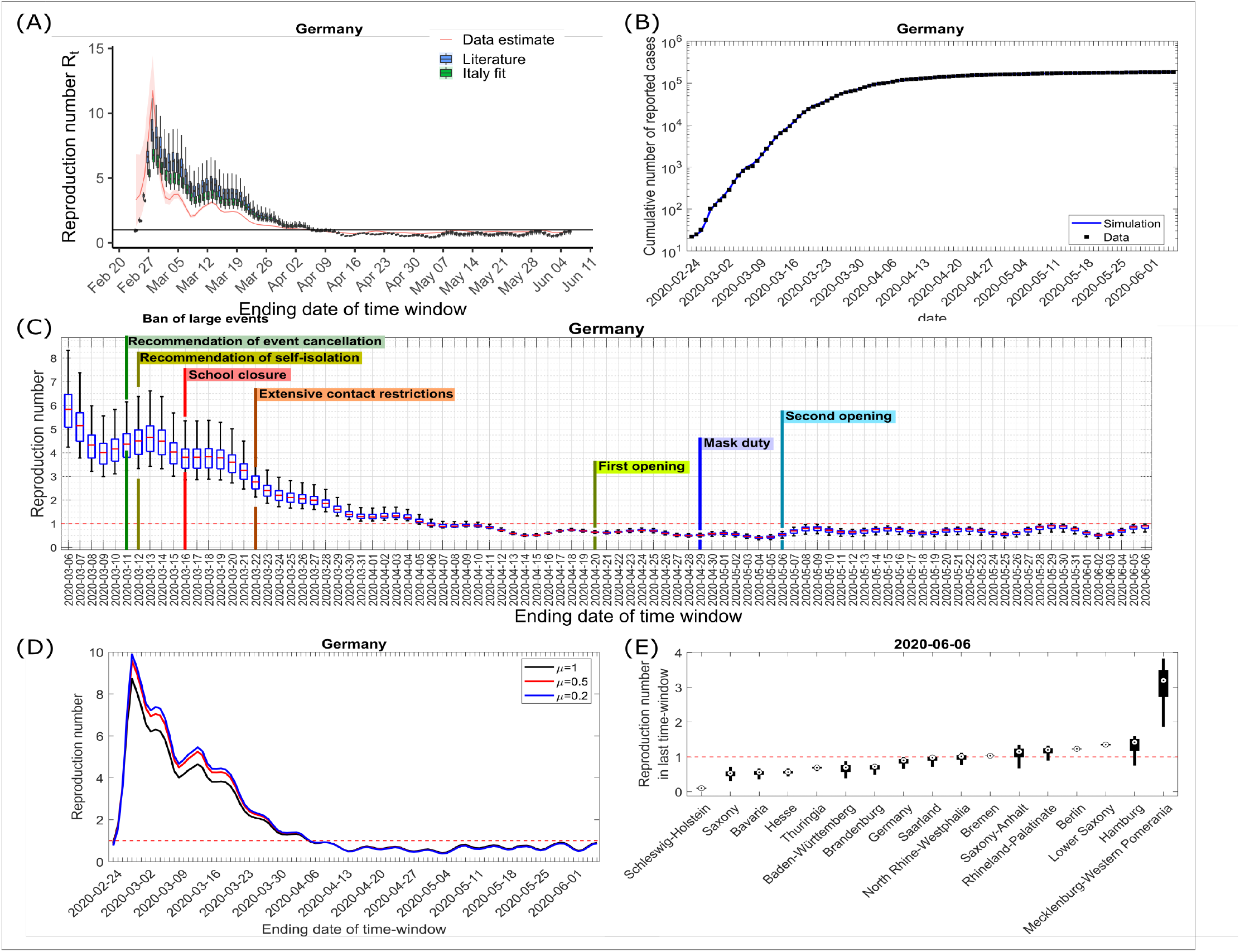
Data for Germany were fitted to the cumulative number of reported cases in a sliding time window with a size of one week. Parameters from Table 1 were used and the transmission rate *R*_1_ was varied (see Methods). (A) Time-varying reproduction number *R*_*t*_ resulting from the fit. The parameter sets were randomly sampled within the ranges in Table 1 and, upon refitting, this induced a variability of reported *R*_*t*_-values. The box plot shows the median, 25- and 75-percentiles as well as the minimum and the maximum values. Both used parameters sets (literature-based and derived from Italy-fit) are compared to the *R*_*t*_-values (red curve) calculated with the publicly available code from Imperial College. (B) The median of fitting results in (A) with literature based parameters is shown for the cumulative number of reported cases and compared with data from [33–35]; own calculation and design. (C) Enlarged result in (A) based on the literature-based parameter set together with the timing of installing and releasing NPIs in Germany. (D) Same analysis as in (A) for different assumptions on the fraction of unreported cases, where *µ* = 1 reflect no unreported case, *µ* = 0.5 means unreported cases are as frequent as reported cases, *µ* = 0.2 assumes 5-times more unreported cases. Only the median value is reported. (E) The same analysis as in (A) was repeated for each federal state in Germany separately (see Supplementary Fig. S3). Here, the last reported *R*_*t*_-value in each federal state of Germany sorted by median values is shown as box plot (same as in (A)). The horizontal line shows *R*_*t*_=1. (A-E): Each data point is a result of 100 randomly sampled parameter sets.

### Basic (*R*_0_) and time-varying (*R*_*t*_) reproduction number

The basic reproduction number *R*_0_ is defined as the expected number of secondary cases produced by a single infection in a completely susceptible population. It can be calculated from the parameters of the respective model [23–26] after fitting the model to data for a given time period during the epidemic. While *R*_0_ provides valuable information on the viral dissemination dynamics in the absence of immunity or awareness of the epidemic, the dynamics of the epidemic over time will be heavily influenced by development of immunity in the population [27], policy changes to minimize infection risk [28], and individual behavior in response to public awareness of a disease [29]. Hence, a practically more useful quantity during an outbreak is the time-dependent reproduction number *R*_*t*_ describing the expected number of secondary cases per index case at a given time of the epidemic. This quantity has to be derived from incidence data over time and reflects the multifactorial impact of NPIs, behavior changes, seasonal effects, etc. on the dynamics of viral spread.

In epidemic models with multiple compartments, *R*_0_ can be derived with the next generation matrix method [25]. The compartments with infected individuals are divided into two contributions with respect to their dynamics: new infections entering the compartments and transfer of infected into and out of the compartment to other compartments. The Jacobian matrices of these two quantities *F* and *V* describe the generation of new infections and the transfer across compartments, respectively [30, 31]. The elements *G*_*ij*_ of *G* = *FV* ^−1^ are related to the expected number of secondary infections in compartment *i* caused by a single infected individual of compartment *j*.

The reproduction number *R*_0_ for the present model is given by the dominant eigenvalue of *G*, i.e.:

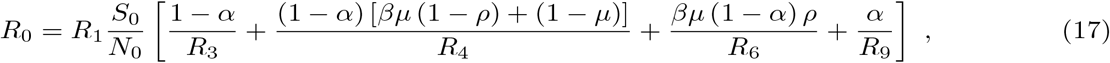

where *N*_0_ is the total population and *S*_0_ is the susceptible population, both at the beginning of the investigation.

For the analysis of *R*_0_, we use the first three days of reported cases as the infected state (*I*_*H*_), and impose them as initial condition for the exposed at one incubation period earlier. This assumption takes into account that the transmission from the first exposed individuals has not happened before the time 1/*R*_2_ and, thus, the first reported cases shall represent independent sources of the virus rather than being the result of transmissions. Given the initial conditions and using the parameter sets in Table 1, the transmission parameter *R*_1_, which mostly contains information on the contact frequency and, thus, best reflects the individual behaviour in the population with respect to social distancing and other measures to minimise the infection risk, is varied in order to optimize the model dynamics to the observed case data. The determined *R*_1_ together with the other model parameter allows to calculate *R*_0_ according to Eq. 17.

In order to assess the impact of political measures onto the development of the reproduction number *R*_*t*_, the cumulative number of registered cases are used. The cumulative case number is compared to the sum of infected individuals and all subsequent states in the model, i.e. with *I*_*H*_ + *I*_*R*_ + *H*_*U*_ + *H*_*R*_ + *U*_*R*_ + *U*_*D*_ + *D* + *R*_*Z*_. A time window of a width of one week is defined starting at the day of the first reported case [32]. This allows to determine *R*(*t*_0_) in the first week and to define proper initial conditions for the first sliding time window (Fig. 2(B)). Then, in repeating cycles, the best *R*_*k*_ := *R*(*t*_*k*_) (with *k* = 1..*N*) for each time window at time *t*_*k*_ is determined by

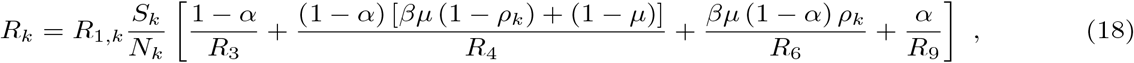

where *ρ*_*k*_ := *ρ*(*t*_*k*_) denotes the average value of the time-varying parameter in the *k*-th time-window. A new set of initial conditions is defined a day later, including the reduced fraction of susceptible individuals *S*_*k*_/*N*_*k*_, with *S*_*k*_ := *S*(*t*_*k*_) and *N*_*k*_ := *N* (*t*_*k*_) the values of susceptible and total population at the starting time *t*_*k*_ of the *k*-th time window. Note that fatal cases reduce the total population. *R*_1,*k*_ := *R*_1_(*t*_*k*_) is determined by fitting to the data in this time window. In cycles the time window is shifted one day later. The series of *R*_*t*_-values for each of the sliding time windows at time *t*_*k*_ is reported at the final date of the time window.

For the prospective study, the state of the model at the last time of *R*_*t*_ evaluation as well as the *R*_*t*_ value itself are kept and used as initial conditions for the model. To assess the impact of different scenarios, the *R*_*t*_-value is subsequently imposed to values of 5/3, 4/3, 1, 2/3, and 1/3 to mimic release, maintenance, or intensification of NPIs, respectively. The cumulative number of infected individuals and the number of occupied ICUs, hospital beds, and deaths is reported. More observables are found at [8].

The distribution of observables and *R*_*t*_-values is generated by reiteration of the analysis under varying model parameters randomly drawn from a uniform distribution within the range provided in Table 1. The box plots in the figures show median, 25- and 75-percentiles as well as minimum and maximum values from these analyses.

## Results

Based on a classical model of infection epidemics, we developed a mathematical model particularly adapted to the requirements and specificities of the COVID-19 outbreak (SECIR-model, Fig. 1). The basic reproduction number is a good measure for the long-term evolution of an epidemic that can be derived from such models (see Methods, Eq. 17). However, it assumes constant conditions over the whole period analysed. In particular, the effect of NPIs on viral spreading enters the reproduction number only in a way mixed with the time before NPIs. For the evaluation of NPI effects on viral spreading, a time-varying reproduction number *R*_*t*_ has to be determined [32]. We opted for a shifting time window (Fig. 2(B)) in each of which *R*_*t*_ is determined and developed an automatized algorithm for the fast analysis of the current *R*_*t*_ (see Methods, Eq. 18). Importantly, each time window is not analysed independently but includes the history of the epidemic by starting from the saved state of the simulation at the beginning of each time window. This analysis was developed for the sake of providing a daily updated evaluation of the reproduction number [8] suitable to support political decisions on NPIs in the course of the COVID-19 outbreak and applied to German federal state data.

The cumulative reported cases are reproduced by the model in each time window, giving rise to a time evolution of the reproductive number *R*_*t*_ (Fig. 3(A,B)). The large initial value at February 28^th^ results from a sudden increase of independent first reported infections, possibly related to people coming back from holiday. This leads to an overshoot of the *R*_*t*_ value in a strength depending on the size of the time window used for analysis. Later on, the nationwide NPIs imposed in Germany included the recommendation for cancellation of large events on March 11^th^, 2020, followed by recommendation of self-isolation issued on March 12^th^, 2020 [36]. Then nationwide school closures on March 16^th^, 2020 [37], and extensive contact restrictions followed on March 22^nd^, 2020 [38] (see Fig. 3 (C)).

The timing of these NPIs suggests that they are responsible for the observed reduction of the reproductive number *R*_*t*_. From there on, the reproduction number went continuously down to a value near 1 as of April 6^th^, 2020. This illustrates that the NPIs imposed appear to have had a strong effect on the dynamics of the COVID-19 epidemic.

NPIs were released in Germany on April 20^th^, 2020 for the first time. Shops were opened, and a few days later, wearing masks became compulsory. The *R*_*t*_-value reacted with a delay of 2-3 weeks and increased by roughly 0.2 and then relaxed again by 0.05, presumably in response to the imposed masks. The second release of measures was decided on May 6^th^ and widely implemented on May 11^th^, 2020 and involved a carefully opening of child care and schools as well as restaurants. However, all of those were opened with imposed contact distancing. Again, three weeks later, the *R*_*t*_-values raised to 0.9 as of June 6^th^, 2020, where it remained since then (not shown). This observation illustrates the sensitivity of the virus to NPIs as well as the possibility to partially release NPIs without losing control of the epidemic, provided the population keeps social distancing and hygiene measures in place. Without the latter, viral spreading would be re-initiated given the basic reproduction number and an only small fraction of immunized individuals.

The number of unregistered cases is not well known in Germany. In the model, we introduced a parameter *µ* that captures which fraction of real cases is actually registered (Fig. 1). In order to demonstrate the importance of the number of undetected cases for the interpretation of the results, we compared the results for *µ* = 1, *µ* = 0.5, and *µ* = 0.2. It turns out that the *R*_*t*_-value derived from a model with unregistered cases is slightly enhanced but remains in the same range (Fig. 3(D)).

Next, the impact of NPIs in the different federal states of Germany was analysed with the same methodology (Supplementary Fig. S3). The early cases in Bavaria, North Rhine-Westphalia, and Baden-Würtemberg can be distinguished from later outbreaks in other federal states. While there is a large diversity of epidemic onset and intermediate developments particular to individual federal states, the overall tendency converges to values below *R*_*t*_ = 1. The coherent reduction of the reproduction number after nationwide implementation of several NPIs together with further measures specifically applied in different federal states speaks for the efficiency of the measures and the responsiveness of the population to the NPIs.

The latest *R*_*t*_ in the different federal states is compared and ranked in Fig. 3(E). Most federal states hit early on by the COVID-19 outbreak exhibited rather low reproduction numbers. The fluctuations, however, are raised by different releases of measures at later time-points. Federal states with overall low case numbers, like Mecklenburg-Vorpommern, exhibit large fluctuations of *R*_*t*_-values.

The model can be used to estimate the dynamics of the load for the health care system. Based on *R*_*t*_-values from Fig. 3(A) we investigated hospitalization and deaths during the epidemic (Fig. 4). The number of deaths and new daily reported cases well captured the trend in the data. The estimated peak (median) of daily new reported cases occurred on March 28^th^ with 6700 cases, 11600 occupied hospital beds on April 5^th^, and 2900 ICU beds on April 8^th^. These numbers stayed within the capacities of the German health care system.

**Fig 4.**
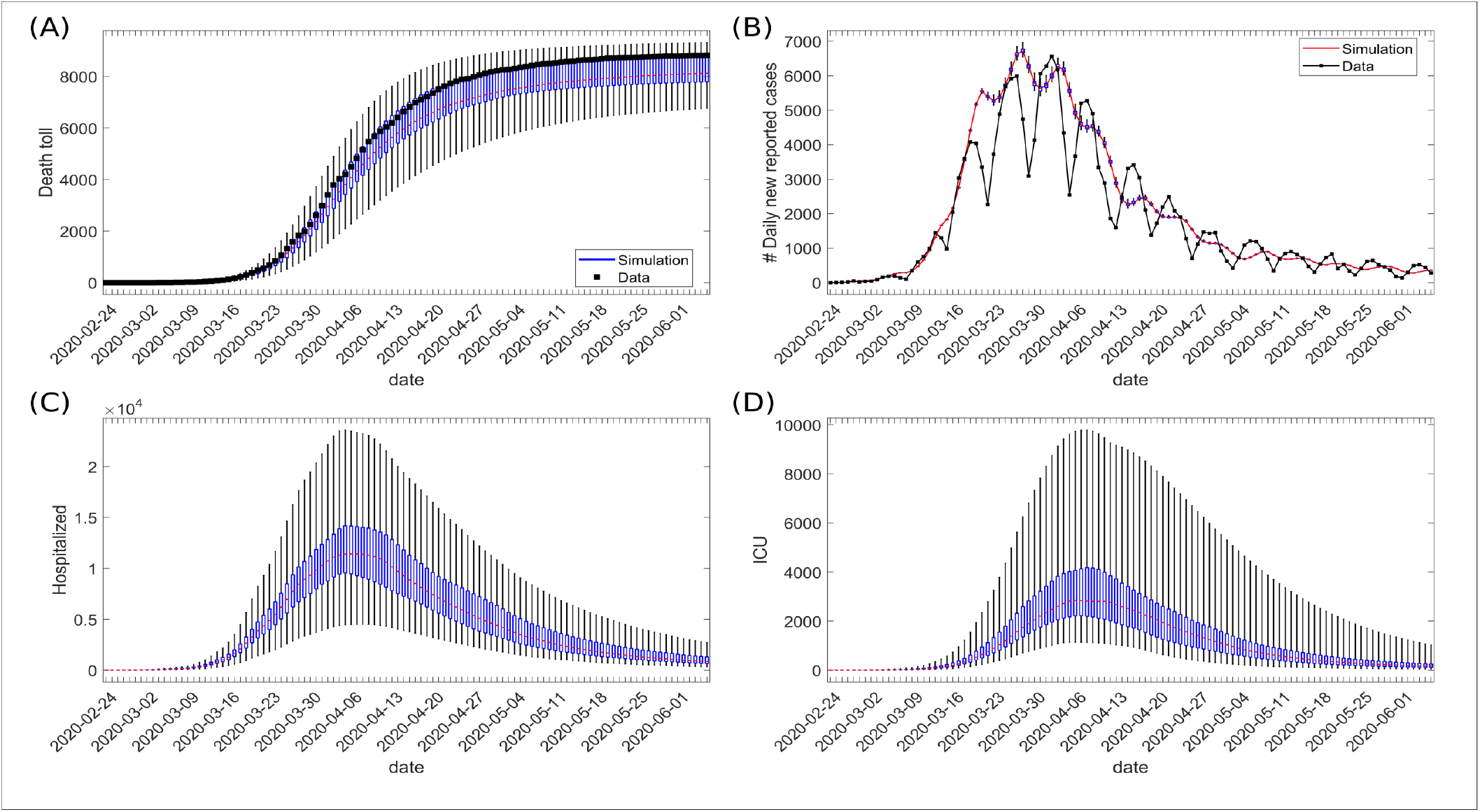
Time-evolution of variables during epidemic. The distribution of simulated values for the last date of sliding time-window is shown for (A) cumulative number of deaths and (B) registered daily new cases, number of hospitalized cases, and (D) number of cases in ICU. The data for analysis were taken from [33–35]; own calculation and design.

We next investigated different prospective scenarios from the final date of the data fitting phase by keeping the state information of the model (Fig. 5(A)). Starting from the last state of the model for Germany (base scenario with *R*_*t*_ ≈ 0.9), thus, including the complex distribution of individuals onto the different compartments of the model at this time, the simulation was continued for one year with the last value of the reproduction number kept for the whole period (Fig. 5(B-F), black). This contained the infection, but was not able to stop it in a short time.

**Fig 5.**
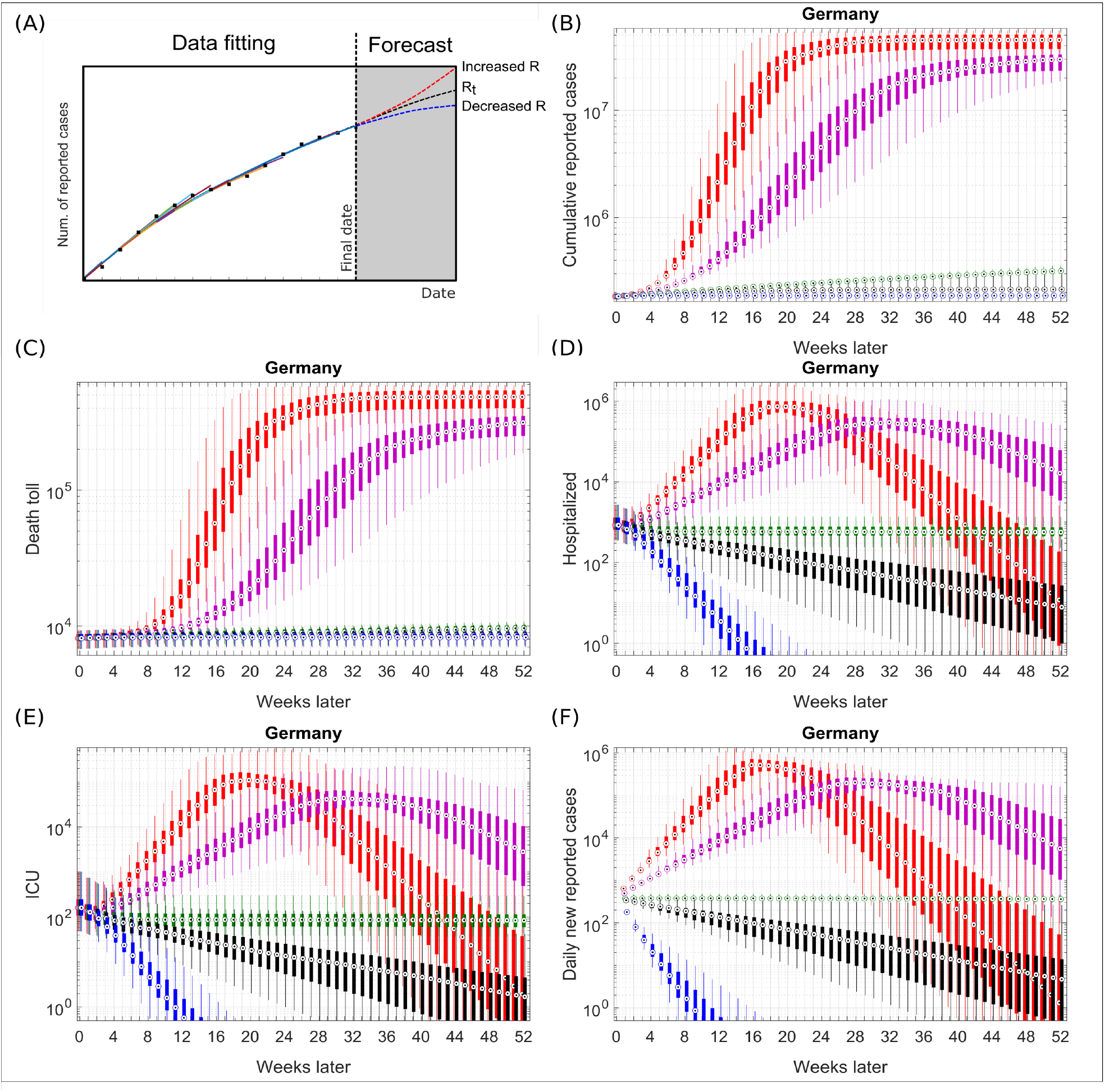
Starting from the final state in Fig. 3(A), a value for the transmission rate *R*_1_ corresponding to *R*_*t*_ values of 5/3, 4/3, 1, the last estimated *R*_*t*_ value, and 1/3 (colors) was assumed for each of the 100 simulations. The simulations were continued for 1 year from this last time point. Box plots show the 25 and 75 percentile as well as min and max. (A) Cumulative reported cases; (B) Hospitalized patients; (C) occupied ICU beds; (D) cumulative deaths. All simulation results are presented on log-scale. Case data before the predicted time from [33–35]; own calculation and design.

In order to assess the impact of releasing NPIs, the reproduction number was set to 1, 4/3 and 5/3 (Fig. 5, green, magenta and red, respectively). A moderate release of NPIs to a degree that induces a reproduction number of 1 induces a controlled continuation of the epidemic (Fig. 5, green). This is associated with an increased number of deaths (by 1100 additional cases compared to the base scenario) and a hundred occupied ICU beds during the whole year.

Any further release of NPIs associated with reproduction numbers larger than 1 leads to a major immunisation of the population (Fig. 5, red and magenta), and to an overwhelmed health care system with a peak in the range of 1 million patients hospitalised and in order of 100,000 ICU beds needed to treat the patients with complications (Fig. 5, red). The value of the peaks increases with increasing the release of the measures, and the time of the peak occurs earlier (Fig. 5(D,E), red versus magenta).

Intensification of NPIs, modelled by a constant reproduction number of 1/3 (Fig. 5, blue), leads to a more efficient suppression of the virus. A complete elimination of the virus, as it appears in the model, is hard to achieve in reality because of open borders to European neighbors and unknown viral properties which might allow it to reappear under particular conditions.

A limited number of daily new cases per day might well be controlled by early detection with extended testing capacities and improved technologies of tracing back contacts together with subsequent preventive isolation of those contacts. Thus, an important question is how long NPIs would have to be kept in place until all new cases can be controlled by public health departments. Assuming 300 detected cases per day to be manageable, we calculated the time needed to achieve this number of daily new cases given different levels of the reproduction number (Fig. 6). Given the reproduction number of June 6^th^, 2020 (Fig. 6(A)), we find that this number would be achieved within less than three weeks for registered cases, and 5 weeks for the infectious cases. The unknown number of undetected cases (*µ* < 1) influences this time. Although the duration to reach 300 new registered cases is comparable for *µ* = 1 and *µ* = 0.2, the infectious population significantly increases with larger fractions of undetected infections (Fig. 6, black versus red).

**Fig 6.**
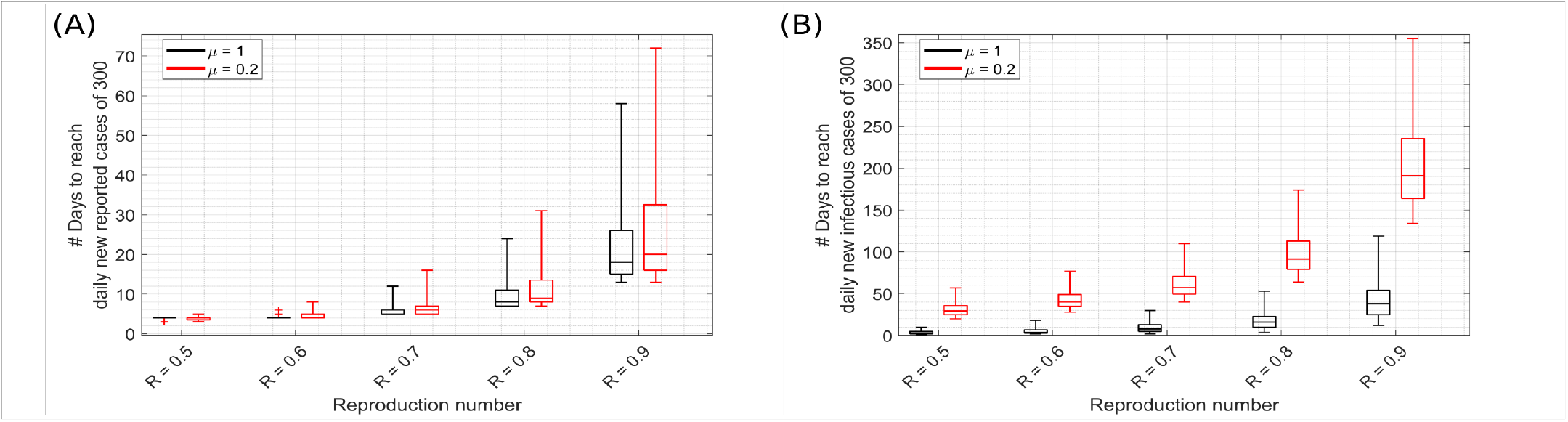
Starting from the last state of the model in Fig. 3(A) for Germany, (A) new reported cases and (B) new infectious cases per day (outflow from exposed compartment, see 1) in whole Germany was estimated in a prospective analysis with different imposed fixed values of reproduction number *R*. For each setting, no (*µ* = 1, black) and five-fold (*µ* = 0.2, red) undetected cases were assumed. Box plots show 75 and 25 percentile and minimum and maximum values.

## Discussion

The SECIR model is a classical mathematical model, which we adapted to the specificities of the recent COVID-19 outbreak. It can capture the qualitative aspects of how the incidence, patients admitted in hospitals and intensive care units as well as deaths alter as days progress during the COVID-19 outbreak. As parameterisation is essential for the quality of the predictions, two reference parameter sets were determined by thorough analysis of the literature on COVID-19 and an independent analysis of Italian data. The results discussed are consistent between both parameter sets, which increases the credibility of the model results.

The development of reported cases in Germany was analyzed and the time-varying reproduction number was estimated. It was found that the reproduction number dropped to values below 1 early in April 2020, likely in response to the NPIs installed mid of March. The projection of the future development of the epidemic can be characterized by three different scenarios:

A. Uncontrolled epidemic with many fatalities and overwhelmed health care system;
B. Long-term ongoing infections treatable with a reasonable health care capacity;
C. Maintenance of NPIs until a controllable low number of incidence is reached.

The scenarios must not be considered as quantitative long-term predictions, but rather should give a qualitative orientation of realistic future developments under certain assumptions.

### Model predictions are structurally limited to a few weeks

The predictive power of the model was analysed by comparing the *R*_*t*_-value as determined by the data and a forecast of daily new cases for the next or the next two weeks (Supplementary Fig. S1A and S2A). Those predictions were compared to the real daily new cases and an optimal *R*_*t*_-value that would predict those best was determined. It turns out that the *R*_*t*_-values from data fitting are near to these optimal values with a tendency to slightly over-estimate the *R*_*t*_-values (Supplementary Fig. S1B and S2B). Larger assumed *R*_*t*_-values led to substantially worse predictions (not shown). This result shows that the model is able to make fair predictions on a scale of a few weeks. However, a good prediction requires a stable situation in the society. The error increases if the population changes behavior or if NPIs are imposed or released. A long-term prediction is structurally impossible because the impact of behavior, NPIs, seasonal effects or viral mutations are not foreseeable based on the available data.

### A complete release of NPIs would still overwhelm the health care system

Releasing NPIs together with returning individual’s behaviour to as before the epidemic will inevitably induce an acceleration of viral spreading in Germany corresponding to scenario A. This holds true as long as neither herd immunity was reached nor other reasons why the susceptible population might be smaller than expected have been identified. In this scenario, the health care system will in expectation need a peak capacity of several 100,000 ICU beds, compared to about 10,000 free ICUs currently available [39]. Many patients in life-threatening conditions would simply not be treated and die. The total number of expected deaths in this scenario is huge and not tolerable.

### It is unrealistic to wait for a major immunisation of the population

The strategy to keep the health care system functional and to delay viral spreading until major parts of the population are immunised, i.e. scenario B, is likely to fail. The delay needed to keep the number of cases within the limits of the health care capacity is too long. The fraction of the immunised population in this scenario is in the range of 1% after one year (0.31% calculated as the ratio of recovered to initial susceptible population on June 6th, 2020). This fraction was not even reached in Hubei province in China (estimated as the ratio of case numbers to total population). Assuming five times more unreported cases and assuming long-term immunity of infected survivors, substantially less than 5% of the population would be immune against the virus (1.6% estimated as the ratio of recovered to initial susceptible with *µ* = 0.2 in the model on June 6th, 2020). Thus, the reduction of transmission probability by immunisation of the population is too small to prevent a renewed outbreak upon release of all containment measures.

While scenario B is possible in terms of rescuing a large part of the population from death by COVID-19 infections, it is damaging many other aspects of the society. It will lead to a major economic burden [4], induce unemployment and other collateral damages. Moreover, social distancing is associated with increased suicide risks [40] and the treatment of other diseases could pose a strong load on the health care system as well [41].

### NPIs can be released to some extent, but the virus is prepared to regrow

The reproduction number was still in a regression phase when the first NPIs were released in Germany on April, 20^th^, 2020. As a consequence, the reproduction number increased again but was kept in a range of 0.75. Thus, the opening of the shops revealed two major lessons learned: (i) The virus is there and will restart spreading if all measures are released; (ii) the society tolerates a certain degree of releasing NPIs without re-entering scenario A or B.

### Responsible behaviour of the population allows for looser NPIs

The stability of the reproduction number despite releases of NPIs has to be considered as the result of a complex co-existence of different effects. Opening the shops was done together with a recommendation of a limitation of the number of people per surface and of avoidance of direct contacts. Indeed, in the UK the reduction of inter-personal contacts was estimated to bring down the reproduction number below one [42]. Face masks were recommended in Germany and compulsory after one week of open shops. Thus, social distancing and the trained new culture of mutual care was interfering with the increased risk of viral transmission. In addition, seasonal effects might have helped to contain the virus further. It is plausible that a major seasonal effect is an increased frequency of being at fresh air and more frequently exchanged air in closed rooms during summer time. Aerosols may accumulate in closed rooms [43, 44] and there is evidence that they contribute to virus transmission [45, 46], suggesting that such seasonal effects could have a substantial effect on the overall epidemic evolution.

## Releasing NPIs delays reaching a controllable number of new infections

NPIs, together with a new culture of inter-personal behavior, were efficient in reducing the reproduction and the daily new case numbers. We showed that a lower reproduction number accelerates reaching a low number of daily new cases in a non-linear way. Thus, increasing the reproduction number by releasing NPIs, even if not re-initiating scenarios B or A, delays the reduction of the daily new cases. In view of the non-linearity of the delay with larger reproduction numbers, one may speculate that it might be more advantageous in terms of health and economy to keep NPIs and the inter-personal culture of distance in place in order to quickly achieve a controllable low number of daily new cases [47]. The model showed that a large number of undetected cases increases the delay in reaching a target number of daily new cases. However, the real number of undetected cases is still not known. Seroprevalence studies suggest undetected cases substantially less than 10-fold of the detected cases in Germany [48, 49].

### At low incidence, NPIs can be widely released

In June 2020, the daily new case number was fluctuating around 300 in Germany. Many of the detected cases were associated with super-spreading events (citation), making it difficult to track all contacts. Still, the low overall incidence number allowed to control the situation by contact tracing and testing combined with quarantine measures to prevent a regrowth of the epidemic. Thus, the proposed strategy to bring down the daily new case numbers to a level that can be controlled by contact tracing and testing appears to be efficient. Additional NPI releases like careful and partial opening of schools from May 6^th^, 2020, did not appear to induce a major effect on the epidemic dynamics.

Established social distancing and hygiene measures together with containment of individual cases before major chains of infections were induced may be considered sufficient to keep the virus under control. A comparable strategy of identification and isolation of infection clusters also worked well in Japan [50], a country with an established culture of inter-personal protection.

### NPIs have to be adapted to the local needs rather than being applied globally

The analysis of the individual federal states in Germany revealed local differences. The same holds true for different regions of Italy, as revealed by the large diversity of model parameters specific for each region. The federal states appear to witness different phases of the outbreak and NPIs exhibit different kinetics of impact. States with lower numbers of reported cases appear less responsive to measures. For that reason, it appears appropriate to fine-tune the NPI measures on a federal or even district level and to adapt to the local situation. The full analysis of all federal states is available at [8] and clearly emphasizes local diversity of the epidemic. A locally more refined modelling may help to inform political decisions in each federal state or district separately.

### A long-term control of the epidemic requires flanking measures

While a complete elimination of the virus is hard to achieve in an open country as Germany in the centre of Europe, mitigation of the epidemic with a low number of remaining new cases per day appears manageable. This requires (i) strong public health departments performing an efficient tracing of contacts of a newly infected person, which might involve support by tracing apps; (ii) substantially extended testing capacities applied to a wide cluster of individuals around an identified new case; (iii) the development of fast testing methods that can prevent outbreaks in vulnerable subpopulations like in resident homes; and (iv) a new societal culture of mutual care and responsibility, alongside increased hygiene possibilities in public places.

## Data Availability

The full data set associated with the research of the paper is made available online. See URL below.

http://secir.theoretical-biology.de/

## Acknowledgments

We are grateful to Werner Hofmann and Alexander Kuhlmann for insightful suggestions. We thank Rebecca Ludwig for support in data acquisition and for revising the manuscript. This project has received funding from the European Union’s Horizon 2020 research and innovation programme under grant agreement No 101003480 and the Initiative and Networking Fund of the Helmholtz Association.

## Supporting information

### Prediction accuracy

**Fig S1.**
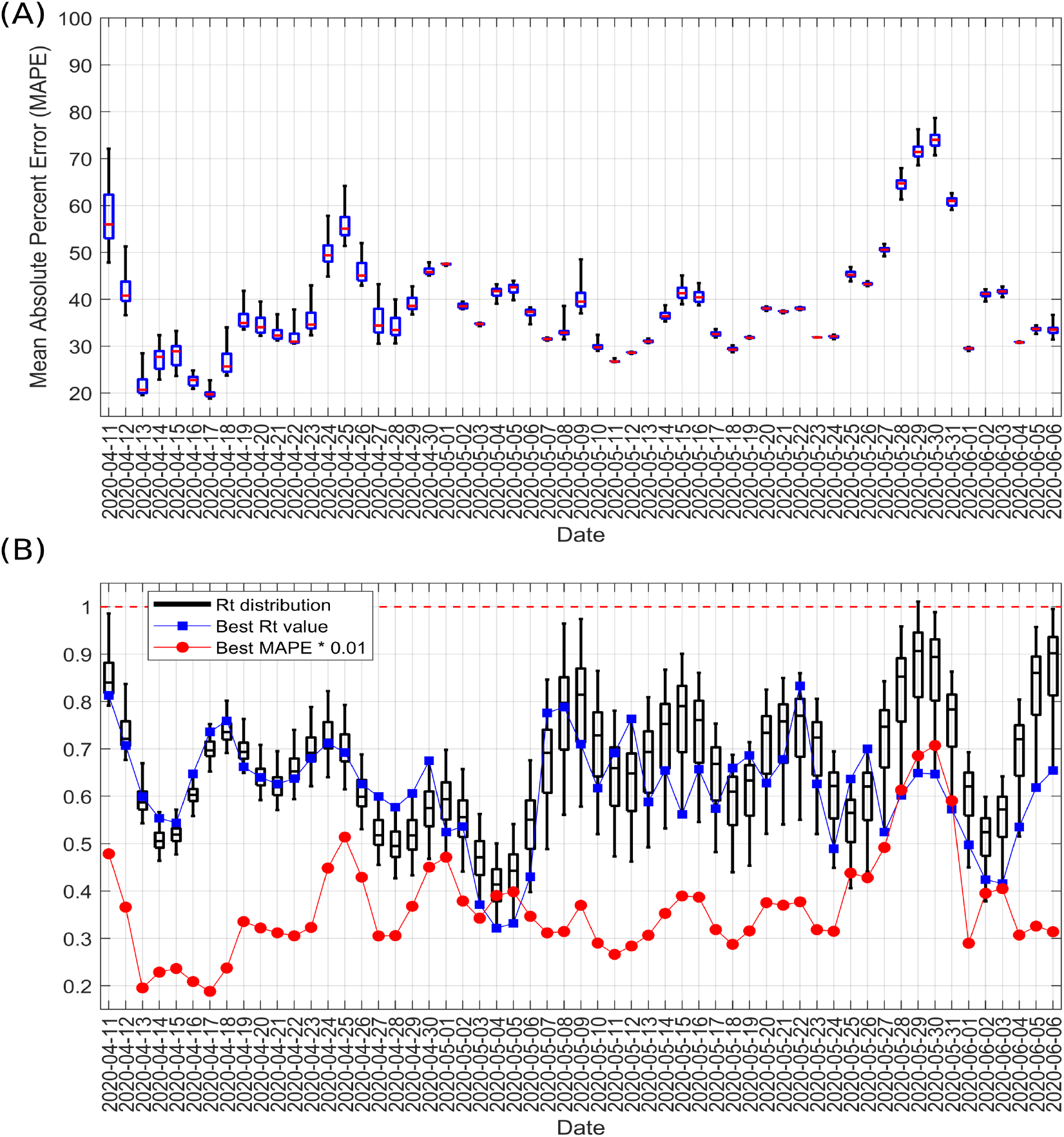
Forecast accuracy. Starting from a given date and model state, the prediction accuracy of the model for the development of daily new reported cases was analyzed for the next 7 days. The values of Rt from 100 simulations as given in Fig. 3A are kept unchanged during this period. (A) The mean absolute percent error (MAPE) of the predictions was calculated for predictions starting from the date on the horizontal axis. The distribution reflects the diversity of model states and corresponding Rt values in the 100 simulations. (B) The Rt-value best predicting the case data in the next 7 days was determined (blue squares) and the corresponding MAPE calculated (red dots). The best Rt-value is compared to the distribution of Rt-values in the 100 simulations underlying the MAPE in panel A. The box plots in panels A and B show median, 25%- and 75%-percentile, minimum and maximum values. Case data was taken from [33–35]; own calculation and design.

**Fig S2.**
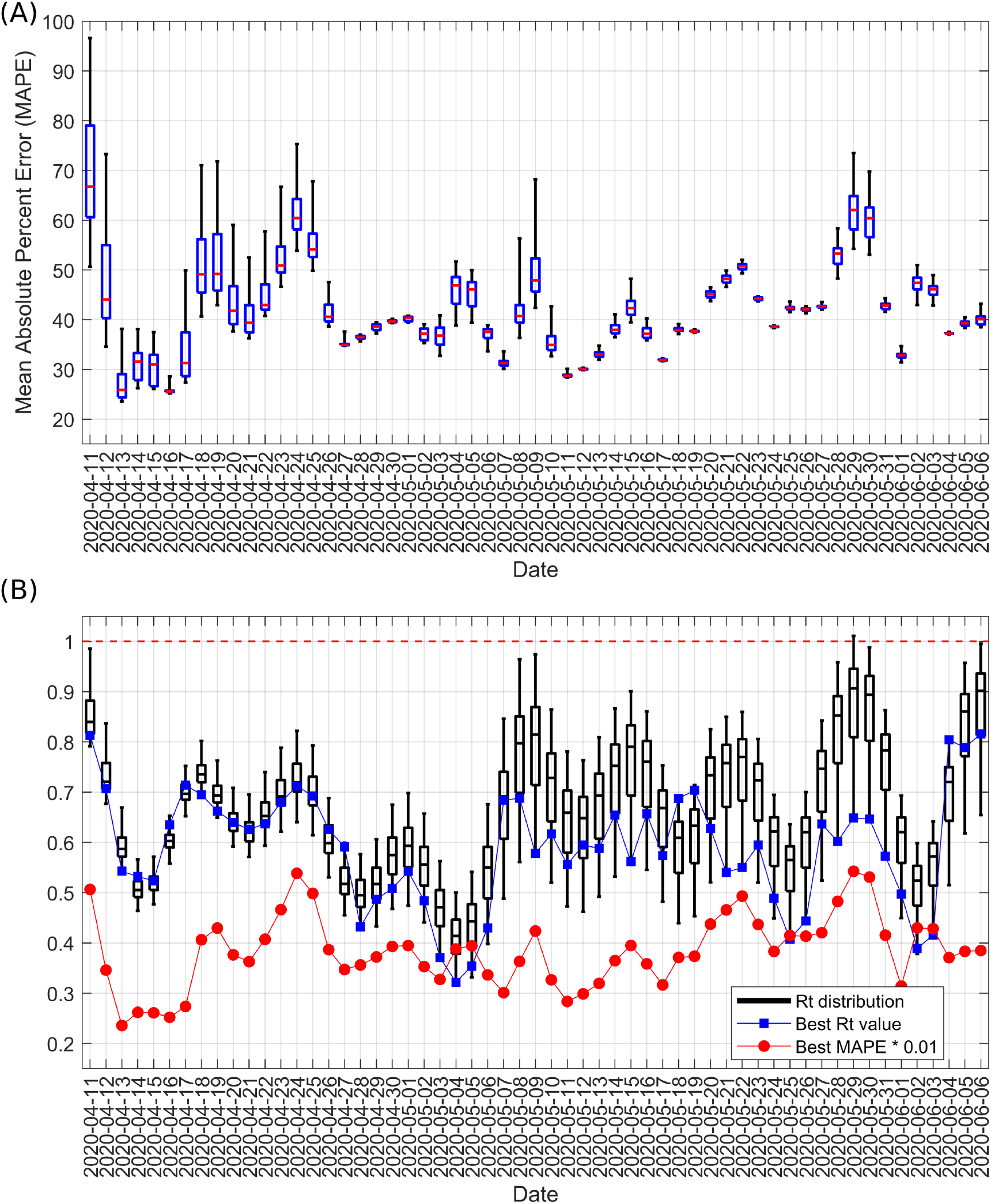
Same analysis as in Fig. S1 but extending the forecast to 14 days.

### *R*_*t*_ in individual federal states

**Fig S3.**
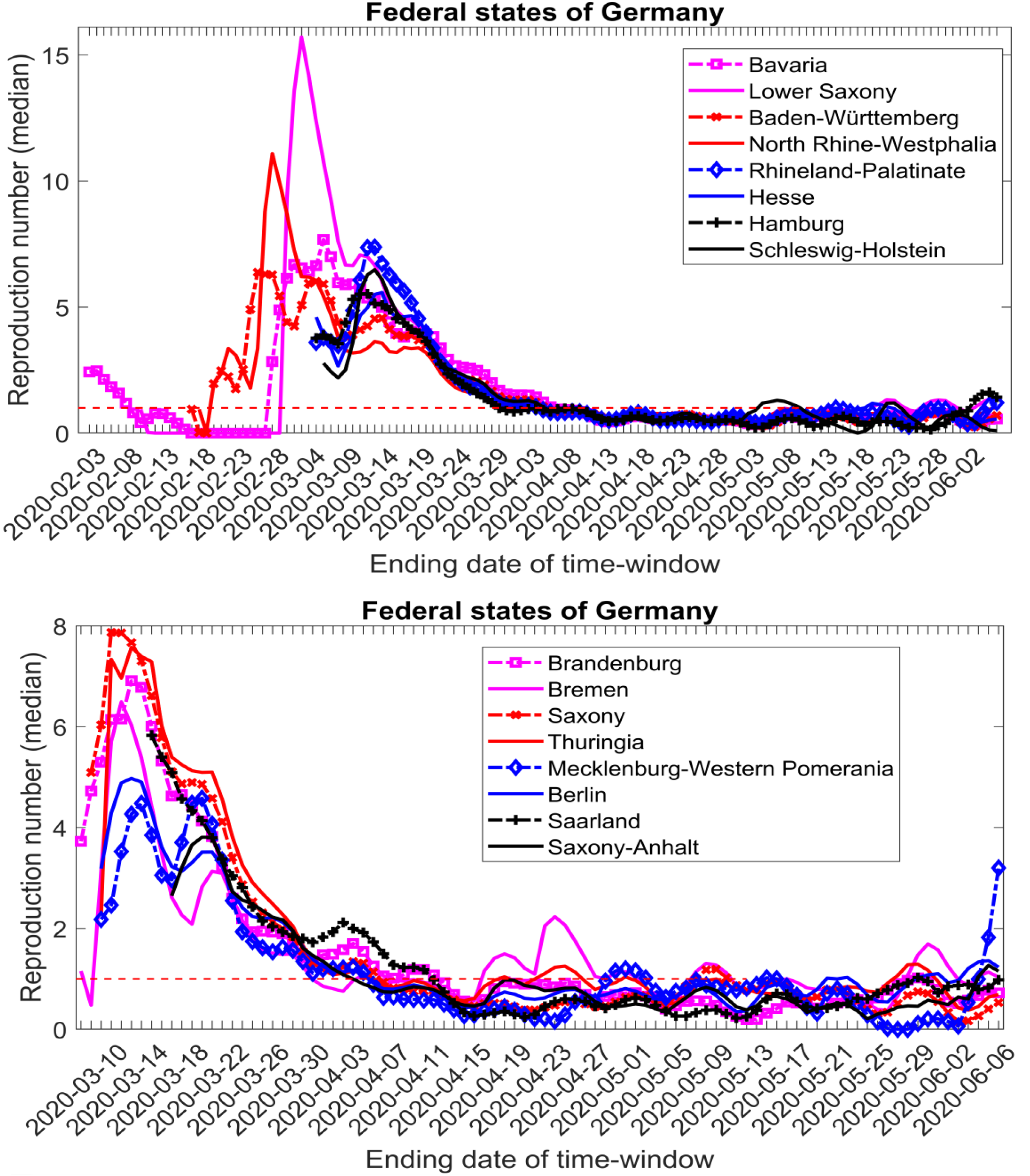
Same analysis of the time-varying reproduction number *R*_*t*_ as in Fig. 3A but for individual federal states separately. For better visualization, only the median values are shown. The states are sorted in the sequence of appearance of first infection cases.

### Parameter description

#### *R*1

R1 represents the product of median contact frequency for a population and the transmission probability of COVID-19 in each of the contacts with a carrier or infected person.

#### *R*2 and *R*3

To estimate *R*2, one needs to know the duration for which an individual remains in a latent non-infectious stage following the transmission of COVID-19 (inverse of this gives *R*2), whereas *R*3 can be estimated as the inverse of the period of being infectious before disease onset. If it is assumed that subsequent infections can occur at random during the infectious period before the onset of symptoms, then the average serial interval will be the sum of the average latent period (from infection to infectiousness) and half the average infectious period before disease onset. Hence, we can state the following equations:

#### Serial Interval

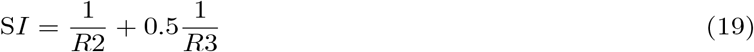

#### Incubation Period

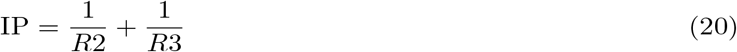

We know that the mean incubation period for COVID-19 is 5.2 days (Li et al. 2020). The mean serial interval is estimated to be 4 days in one study from a total of 28 infector-infectee pairs [5] and 4.4 days in another study based on data from 21 infection chains in Hong Kong [14]. For our calculation, we have used the mean of these two studies as our average serial interval, which is 4.2 days. Using the above equations for the serial interval and incubation period, we have calculated the values of 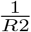 and 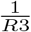, which provide us with a median value of 2 days for 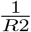 and 3.2 days for 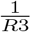. Further assuming that a person once exposed to the infection, spends at least one day in latent non-infectious period and overall, the latent non-infectious period 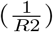 is not greater than latent infectious period 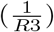, we get a range of 2.6 to 4.2 days for 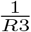. To calculate the corresponding value of 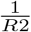, we make use of the median incubation period 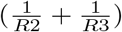 of 5.2 days.

#### *R*4

The inverse of *R*4 is the duration for which the infected individuals with mild symptoms and not requiring hospitalization, remain infectious after their disease onset. To estimate this, we have made use of one study with nine young patients with no underlying health conditions, where the excretion dynamics of reproductive viruses [15] from samples of the throat and sputum were examined. This study suggests active virus replication in the upper respiratory tract in the earlier phase of the disease following onset of symptoms. RT-PCR tests result in detectable viral sub-genomic messenger RNAs (sgRNA) in swabs from throat in the first 5 days after symptoms onset (In Figure 1 (d) in [15], the throat swab cultures are positive up to the 4^th^ day, which the authors mark as sample of 4/5 days). However, we note that active virus is found in the sputum until day 8 for these mildly ill cases. Accounting for some variations, we assume that patients with minor illnesses not requiring hospitalization are infectious for 6 days following disease onset, which results in a value of 6 days for 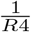. For such individuals, to calculate the median value of *R*4, we neglect a potentially longer period of possible infections transmitted via the sputum, this mode of transmission being especially meaningful in a hospital setting. For our analysis, we have considered a range of 4 – 14 days for 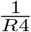.

#### *R*5

The inverse of *R*5 depicts the duration for which the hospitalized patients not requiring further intensive care remain under general hospital care before getting discharge. The median value of 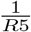 depends on the age structure of the affected people in a particular region. For example, in the case of Braunschweig, the second largest city in Lower Saxony, where 46.35% of the population is under the age of 40, the median value of 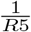 will be biased towards the value of 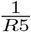 corresponding to this young group. In a Chinese case study [16], the median time for the time span of hospitalization is reported to be around 10 days for the mild cases. Although the WHO-China joint mission on COVID-19 outbreak [17] has reported a median period of 14 days for the hospitalized cases not requiring intensive care treatment, we assumed a period of 10 days to be the median value of 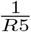 for Germany by considering the age-distribution of infected population here. However, we do vary 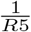 in a wide range of 7 – 16 days while performing the analysis.

#### *R*6

The inverse of *R*6 denotes the time a patient with mild symptoms spends at home before hospital admission due to worsening of the disease condition. We assume that the patients are admitted to the hospital following the onset pneumonia and/or shortness of breath. One Chinese case series [19] reports a median duration of 4 days as the time span that leads to pneumonia in case of COVID-19 following manifestation of disease symptoms. Another study [16] finds the median duration from onset of symptoms to onset of breathing difficulty to be 5 days. A third Chinese case series [18] based on 298 patients admitted to one hospital in Shenzhen has reported that the median time span from disease onset to hospital admission was 5 days. To consider age dependence of 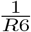, we have assumed that aged patients develop dyspnoea and pneumonia faster than the younger ones, thereby requiring admission to hospitals faster following onset of disease symptoms. To account for such a scenario, we have considered a range of 2.5 – 7 days for 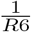.

#### *R*7

Inverse of *R*7 represents the time span spent following hospitalization to admission in an intensive care unit, primarily due to acute respiratory distress syndrome (ARDS). The median value of 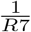 depends on the age structure of the affected people in a particular region. For example, in the case of Braunschweig, the median value of 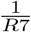 will be biased towards the value of 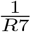 for the age group <40. A Chinese case series [16] has reported the median time span from hospitalization to admission in intensive care units to be around 1 day, although the range varies between 0 – 3 days (IQR) depending on age. The time span spent during hospitalization to admission in ICU is likely shorter as the patients get older. For Germany where the majority of reported infections are in middle-aged groups, we assume the median value of 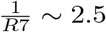 days [12].

As we do not consider the time patients spend in ward beds after having recovered in ICU with an extra compartment, the total duration of the hospital stay

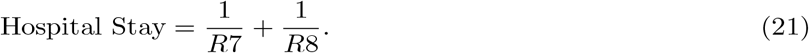

and, thus, the occupancy of hospitals, would be underestimated. WHO-China joint mission on COVID-19 outbreak [17] reports that the total duration of being in hospital for the severely ill patients can be around 3 to 6 weeks. As we were more interested in occupancy than in the exact timing of ICU peaks, we opted to increase the time 1/*R*7 to a range of 4-14 days. This delays ICU occupancy and implies that we must not interpret the exact timing of ICU occupancy and death tolls in this model.

#### *R*8

The inverse of *R*8 depicts the time span spent in ICU before discharge. We vary 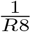 in a range of 5 – 16 days. This is consistent with the median value of 9 days reported by RKI [12].

#### *R*9

The inverse of *R*9 is primarily the duration for which the asymptomatic infected individuals remain infectious following their latent non-infectious period. As these individuals do not show symptoms, we assume that they remain infectious for a shorter time as compared to those who develop even milder symptoms. From the aforementioned discussion, we note that the cases with mild symptoms remain infectious for a period of (1/R3 + 1/R4). Hence, our assumption restricts 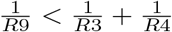. If we further assume that asymptomatic people are following a similar trajectory as the people with mild symptoms, and randomly become non-infectious during the whole duration of 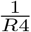, this results in a median value of 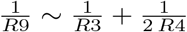. We use the same formulation while varying R3 and R4 in ranges as described above.

#### *R*10

The inverse of *R*10 denotes the time span a patient admitted in ICU spends there before dying. It is estimated from time to death from onset of symptoms, which is reported to be around 14 days [20]. Hence, the median value of 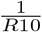 is calculated using the following estimate:

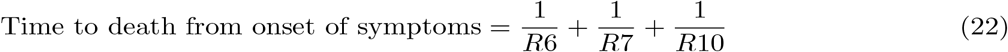

This gives: 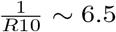 days as an average estimate (considering 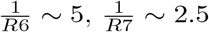). For a city having demography with a predominantly aged population (70+), the median time span to death from onset of symptoms can be around 11.5 days [20]. We have varied 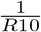 over a range of 3.5 – 7.5 days.

#### *ρ* (Fraction 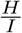

It might be difficult to calculate from the earlier Chinese case studies because even people with non-severe courses of disease were admitted to hospitals for isolation [12]. Rather, one may estimate this by asking in what proportion symptomatic cases were severe. One Chinese case series estimated it to be around 19% [21]. The estimates for UK data [22, 51] have been age specific, and this fraction can be up to 27.3% depending on age. USA data [52] suggests about 12% of the confirmed cases were in hospital till March 16, 2020 but it is important to note that this as well will not reflect the true fraction of symptomatic cases requiring hospitalization. Diversity in hospitalization conditions across regions and countries make it difficult to estimate a true average of this fraction, prompting us to assume it to be around 20% as an estimate for overall population, but keeping the room to alter it within realistic range while fitting the region specific data. We also assume that this fraction increases with age. We have performed our analysis over a range of 0.10 – 0.35 for *ρ*.

#### *ϑ* (Fraction 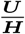)

In one Chinese case series of 138 patients [16], the percentage of hospitalized patients requiring intensive care support was 26%. We assume that this fraction increases with age. We vary *ϑ* within a range of 0.15 – 0.40.

#### α

Currently, there is no reliable data available about the asymptomatic cases. However, we note that about 9% of the confirmed infectious population in Italy remain asymptomatic [1]. We can also have an idea about this fraction from the manifestation index [12]. The manifestation index describes the proportion of those infected who actually fall ill. Three studies from different settings (cruise ship outbreak, evacuated returning travellers, contact-based case search) gave figures of 51% [53], 69% [54] and 81% [55]. Hence, in a scenario where extensive tests are not performed and contacts are not traced properly, this fraction could be higher. However, in a setting where contacts have been traced properly, thereby effectively isolating the exposed population in an early stage before they become asymptomatic carriers, and when extensive tests are performed, this fraction would be minimal. As a median value, we assume it to be nearly 9%. We further assume that most of the people will show up symptoms as age increases. We vary *α* for a range of values from 0.01 – 0.5.

#### β

It represents the risk of infection from the infected symptomatic patients as well as captures the risk from all those who are not yet effectively isolated. Therefore, this fraction varies within countries, cultures and healthcare systems. We assume it to be in the range of 0.05 - 1.0. In the best-case scenario, this fraction would be zero, which is too unrealistic due to delay in isolating patients. The maximum value of this fraction is 1, which is also unrealistic as it would mean that even after symptoms occur, every patient is free to make contacts with others as before.

#### *δ* (Fraction 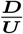)

Mortality rate is often overestimated during an ongoing outbreak, and it largely depends on the healthcare system. In a setting where the healthcare system is overwhelmed, this will be higher. In an ideal healthcare system where we have enough supply of resources (e.g. ICUs, hospital beds), it can be assumed that patients only die after being admitted in ICUs, and thereby can be estimated using 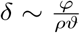; where *ϕ* is the overall mortality. The mortality percentage in case studies is largely age dependent [16, 21] and increases with age. This estimation gives 0.77 as the median value, which we note is towards the higher end, but interpretable as many severely ill patients in ICUs may die from the illness. For our analysis, we have varied *δ* over 0.15 – 0.77.

## Notes

### Competing Interest Statement

The authors have declared no competing interest.

### Funding Statement

This project has received funding from the European Union Horizon 2020 research and innovation programme under grant agreement No 101003480 and the Initiative and Networking Fund of the Helmholtz Association.

## References

1. Istituto Superiore di Sanità. Report sulle caratteristiche dei pazienti deceduti positivi a COVID-19 in Italia. Ilpresente report è basato sui dati aggiornati al 20 Marzo 2020.; 2020. Available from: https://www.epicentro.iss.it/coronavirus/sars-cov-2-sorveglianza-dati

2. an der Heiden M, Buchholz U. Modellierung von Beispielszenarien der SARS-CoV-2-Epidemie 2020 in Deutschland; 2020. Available from: https://www.rki.de/{DE}/Content/{InfAZ}/N/{Neuartiges_Coronavirus}/{Modellierung_Deutschland}.html

3. Hartl T, Wälde K, Weber E. Measuring the impact of the German public shutdown on the spread of Covid-19. Covid Economics. 2020;1:25–32.

4. Dorn F, Fuest C, Marcell G, Krolage C, Lautenbacher S, Link S, et al. Die volkswirtschaftlichen Kosten des Corona-Shutdown für Deutschland: Eine Szenarienrechnung. ifo Schnelldienst. 2020;73(04).

5. Nishiura H, Linton NM, Akhmetzhanov AR. Serial interval of novel coronavirus (COVID-19) infections. Int J Infect Dis. 2020;93:284–286. doi:10.1016/j.ijid.2020.02.060.

6. Flaxman S, Mishra S, Gandy A, Unwin HJT, Coupland H, Mellan TA, et al. Estimating the number of infections and the impact of non-pharmaceutical interventions on COVID-19 in 11 European countries.; 2020. Available from: https://www.imperial.ac.uk/mrc-global-infectious-disease-analysis/covid-19/report-13-europe-npi-impact/

7. Flaxman S, Mishra S, Gandy A, Unwin H, Mellan T, Coupland H, et al. Estimating the effects of non-pharmaceutical interventions on COVID-19 in Europe. Nature. 2020;. doi:10.1038/s41586-020-2405-7.

8. Systems Immunology Group. Complete and up-to-date analysis of Germany and all federal states.; 2020. Available from: http://secir.theoretical-biology.de

9. Robert Koch Institut. Corona Virus Disease 2019 (COVID-19) Daily Situation Report of the Robert Koch Institute.; 2020. Available from: https://www.rki.de/{DE}/Content/{InfAZ}/N/{Neuartiges_Coronavirus}/Situationsberichte/2020-06-17-en.pdf?blob={publicationFile}

10. Vanella P, Wiessner C, Holz A, Krause G, Moehl A, Wiegel S, et al. The role of age distribution, time lag between reporting and death and healthcare system capacity on case fatality estimates of COVID-19. medRxiv. 2020;. doi:10.1101/2020.05.16.20104117.

11. Davies NG, Klepac P, Liu Y, Prem K, Jit M, working group CC, et al. Age-dependent effects in the transmission and control of COVID-19 epidemics. medRxiv. 2020;. doi:10.1101/2020.03.24.20043018.

12. Robert Koch Institut. Steckbrief zur Coronavirus-Krankheit-2019 (COVID-19); 2020. Available from: https://www.rki.de/{DE}/Content/{InfAZ}/N/{Neuartiges_Coronavirus}/Steckbrief.html

13. Li Q, Guan X, Wu P, Wang X, Zhou L, Tong Y, et al. Early Transmission Dynamics in Wuhan, China, of Novel Coronavirus-Infected Pneumonia. N Engl J Med. 2020;382(13):1199–1207. doi:10.1056/NEJMoa2001316.

14. Zhao S, Gao D, Zhuang Z, Chong M, Cai Y, Ran J, et al. Estimating the serial interval of the novel coronavirus disease (COVID-19): A statistical analysis using the public data in Hong Kong from January 16 to February 15, 2020. medRxiv. 2020;. doi:10.1101/2020.02.21.20026559.

15. Woelfel R, Corman V, Guggemos W, Seilmaier M, Zange S, Mueller M, et al. Clinical presentation and virological assessment of hospitalized cases of coronavirus disease 2019 in a travel-associated transmission cluster. medRxiv. 2020;. doi:10.1101/2020.03.05.20030502.

16. Wang D, Hu B, Hu C, Zhu F, Liu X, Zhang J, et al. Clinical Characteristics of 138 Hospitalized Patients With 2019 Novel Coronavirus-Infected Pneumonia in Wuhan, China. JAMA. 2020;. doi:10.1001/jama.2020.1585.

17. World Health Organization. Report of the WHO-China Joint Mission on Coronavirus Disease 2019 (COVID-19). Geneva:; 2020.

18. Cai Q, Huang D, Ou P, Yu H, Zhu Z, Xia Z, et al. 2019-nCoV Pneumonia in a Normal Work Infectious Diseases Hospital Besides Hubei Province, China. SSRN Journal. 2020;. doi:10.2139/ssrn.3542163.

19. Guan WJ, Ni ZY, Hu Y, Liang WH, Ou CQ, He JX, et al. Clinical characteristics of coronavirus disease 2019 in China. N Engl J Med. 2020;. doi:10.1056/NEJMoa2002032.

20. Wang W, Tang J, Wei F. Updated understanding of the outbreak of 2019 novel coronavirus (2019-nCoV) in Wuhan, China. J Med Virol. 2020;92(4):441–447. doi:10.1002/jmv.25689.

21. Novel Coronavirus Pneumonia Emergency Response Epidemiology Team. The epidemiological characteristics of an outbreak of 2019 novel coronavirus diseases (COVID-19) in China. Zhonghua Liu Xing Bing Xue Za Zhi. 2020;41(2):145–151. doi:10.3760/cma.j.issn.0254-6450.2020.02.003.

22. Verity R, Okell LC, Dorigatti I, Winskill P, Whittaker C, Imai N, et al. Estimates of the severity of COVID-19 disease. medRxiv. 2020;. doi:10.1101/2020.03.09.20033357.

23. Wallinga J, Lipsitch M. How generation intervals shape the relationship between growth rates and reproductive numbers. Proc Biol Sci. 2007;274(1609):599–604. doi:10.1098/rspb.2006.3754.

24. Heffernan JM, Smith RJ, Wahl LM. Perspectives on the basic reproductive ratio. J R Soc Interface. 2005;2(4):281–293. doi:10.1098/rsif.2005.0042.

25. Diekmann O, Heesterbeek JA, Metz JA. On the definition and the computation of the basic reproduction ratio R0 in models for infectious diseases in heterogeneous populations. J Math Biol. 1990;28(4):365–382. doi:10.1007/BF00178324.

26. Diekmann O, Heesterbeek JAP, Roberts MG. The construction of next-generation matrices for compartmental epidemic models. J R Soc Interface. 2010;7(47):873–885. doi:10.1098/rsif.2009.0386.

27. Fine P, Eames K, Heymann DL. “Herd immunity”: a rough guide. Clin Infect Dis. 2011;52(7):911–916. doi:10.1093/cid/cir007.

28. OECD. Coronavirus: The world economy at risk.; 2020. Available from: https://www.oecd.org/berlin/publikationen/Interim-Economic-Assessment-2-March-2020.pdf

29. Weston D, Hauck K, Amlôt R. Infection prevention behaviour and infectious disease modelling: a review of the literature and recommendations for the future. BMC Public Health. 2018;18(1):336. doi:10.1186/s12889-018-5223-1.

30. van den Driessche P, Watmough J. Reproduction numbers and sub-threshold endemic equilibria for compartmental models of disease transmission. Math Biosci. 2002;180:29–48. doi:10.1016/S0025-5564(02)00108-6.

31. van den Driessche P, Watmough J. Further notes on the basic reproduction number. In: Brauer F, van den Driessche P, Wu J, Morel JM, Takens F, Teissier B, editors. Mathematical Epidemiology. vol. 1945 of Lecture notes in mathematics. Berlin, Heidelberg: Springer Berlin Heidelberg; 2008. p. 159–178. Available from: http://link.springer.com/10.1007/978-3-540-78911-6_6

32. Cori A, Ferguson NM, Fraser C, Cauchemez S. A new framework and software to estimate time-varying reproduction numbers during epidemics. Am J Epidemiol. 2013;178(9):1505–1512. doi:10.1093/aje/kwt133.

33. Nationale Plattform für geographische Daten. RKI COVID19; 2020. Available from: https://npgeo-corona-npgeo-de.hub.arcgis.com/datasets/dd4580c810204019a7b8eb3e0b329dd6_0/data

34. GENESIS Online. Bevölkerung: Deutschland, Stichtag, Altersjahre.; 2020. Available from: https://www-genesis.destatis.de

35. GENESIS Online. Bevölkerung: Bundesländer, Stichtag, Altersjahre.; 2020. Available from: https://www-genesis.destatis.de

36. Bundesministerium für Gesundheit. Chronik zum Coronavirus SARS-CoV-2 - Maßnahmen des BMG; 2020. Available from: https://www.bundesgesundheitsministerium.de/coronavirus/chronik-coronavirus.html

37. Tagesschau. Corona-Krise: Wo bleiben Schulen zu - und wie lange? - tagesschau.de; 2020. Available from: https://www.tagesschau.de/inland/corona-schulschliessungen-103.html

38. Bundesregierung. Besprechung der Bundeskanzlerin mit den Regierungschefinnen und Regierungschefs der Länder; 2020. Available from: https://www.bundesregierung.de/breg-de/themen/coronavirus/besprechung-der-bundeskanzlerin-mit-den-regierungschefinnen-und-regierungschefs-der-laender-1733248

39. DIVI-IntensivRegister. DIVI IntensivRegister Tagesreport 04.04.2020.; 2020. Available from: https://www.divi.de/images/Dokumente/{DIVI}-{IntensivRegister_Tagesreport_2020_04_04}.pdf

40. Ruiz-Perez I, Rodriguez-Barranco M, Rojas-Garcia A, Mendoza-Garcia O. Economic crisis and suicides in Spain. Socio-demographic and regional variability. Eur J Health Econ. 2017;18(3):313–320. doi:10.1007/s10198-016-0774-5.

41. Bartlett DL, Howe JR, Chang G, Crago A, Hogg M, Karakousis G, et al. Management of Cancer Surgery Cases During the COVID-19 Pandemic: Considerations. Ann Surg Oncol. 2020;27(6):1717–1720. doi:10.1245/s10434-020-08461-2.

42. Jarvis CI, Van Zandvoort K, Gimma A, Prem K, Klepac P, Rubin GJ, et al. Quantifying the impact of physical distance measures on the transmission of COVID-19 in the UK. BMC Med. 2020;18(1):124. doi:10.1186/s12916-020-01597-8.

43. Morawska L, Cao J. Airborne transmission of SARS-CoV-2: The world should face the reality. Environ Int. 2020;139:105730. doi:10.1016/j.envint.2020.105730.

44. Setti L, Passarini F, De Gennaro G, Barbieri P, Perrone MG, Borelli M, et al. Airborne Transmission Route of COVID-19: Why 2 Meters/6 Feet of Inter-Personal Distance Could Not Be Enough. Int J Environ Res Public Health. 2020;17(8). doi:10.3390/ijerph17082932.

45. Jones RM, Brosseau LM. Aerosol transmission of infectious disease. J Occup Environ Med. 2015;57(5):501–508. doi:10.1097/JOM.0000000000000448.

46. Asadi S, Wexler AS, Cappa CD, Barreda S, Bouvier NM D RW. Aerosol emission and superemission during human speech increase with voice loudness. Sci Rep. 2019;9:2348. doi:10.1038/s41598-019-38808-z.

47. Dorn F, Khailaie S, Stöckli M, Binder S, Lange B, Peichl A, et al. Das gemeinsame Interesse von Gesundheit und Wirtschaft: Eine Szenarienrechnung zur Eindämmung der Corona-Pandemie.; 2020. Available from: https://www.ifo.de/en/publikationen/2020/article-journal/das-gemeinsame-interesse-von-gesundheit-und-wirtschaft

48. Streeck H, Schulte B, Kuemmerer B, Richter E, Hoeller T, Fuhrmann C, et al. Infection fatality rate of SARS-CoV-2 infection in a German community with a super-spreading event. medRxiv. 2020;. doi:10.1101/2020.05.04.20090076.

49. Korth J, Wilde B, Dolff S, Anastasiou OE, Krawczyk A, Jahn M, et al. SARS-CoV-2-specific antibody detection in healthcare workers in Germany with direct contact to COVID-19 patients. J Clin Virol. 2020;128:104437.

50. Iwasaki A, Grubaugh ND. Why does Japan have so few cases of COVID-19? EMBO Mol Med. 2020;12(5):e12481. doi:10.15252/emmm.202012481.

51. Ferguson NM, Laydon D, Nedjati-Gilani G, Imai N, Ainslie K, Baguelin M, et al. Impact of non-pharmaceutical interventions (NPIs) to reduce COVID-19 mortality and healthcare demand. Imperial College London. 2020;.

52. CDC COVID-19 Response Team. Severe Outcomes Among Patients with Coronavirus Disease 2019 (COVID-19) - United States, February 12-March 16, 2020. MMWR Morb Mortal Wkly Rep. 2020;69(12):343–346. doi:10.15585/mmwr.mm6912e2.

53. Mizumoto K, Kagaya K, Zarebski A, Chowell G. Estimating the asymptomatic proportion of coronavirus disease 2019 (COVID-19) cases on board the Diamond Princess cruise ship, Yokohama, Japan, 2020. Euro Surveill. 2020;25(10). doi:10.2807/1560-7917.ES.2020.25.10.2000180.

54. Nishiura H, Kobayashi T, Miyama T, Suzuki A, Jung SM, Hayashi K, et al. Estimation of the asymptomatic ratio of novel coronavirus infections (COVID-19). Int J Infect Dis. 2020;94:154–155. doi:10.1016/j.ijid.2020.03.020.

55. Bi Q, Wu Y, Mei S, Ye C, Zou X, Zhang Z, et al. Epidemiology and Transmission of COVID-19 in Shenzhen China: Analysis of 391 cases and 1,286 of their close contacts. medRxiv. 2020;. doi:10.1101/2020.03.03.20028423.

